## Supplemental Table 1 for "A systematic review of the clinical profile of patients with bubonic plague and the outcome measures used in research settings"

**S2a – Risk of bias assessment for case reports**

| **Study title**  ***First author, Year*** | **Demographic characteristics** | **Patient history** | **Current clinical condition** | **Diagnostic tests/assessment** | **Intervention/ treatment** | **Post-intervention clinical condition** | **Adverse/ unanticipated events** | **Takeaway lessons** |
| --- | --- | --- | --- | --- | --- | --- | --- | --- |
| Two Cases Of Bubonic Plague Occurring On Board Ship  *Barnett H. N., 1902* | No | No | Yes | No | No | No | Unclear | Yes |
| A case of bubonic plague on a vessel arriving in the Mersey  *NK, 1905* | No | No | Yes | No | N/A | N/A | N/A | Yes |
| Subacute plague in man due to ground squirrel infection  *McCoy G. W., 1909* | Yes | Yes | Yes | Yes | No | Yes | Yes | Yes |
| Three Cases of Bubonic Plague Arising In England  *Rendle-Short A., 1916* | No | No | Yes | No | No | No | No | Yes |
| Plague -- New Mexico  *United States Centres for Disease Control and Prevention, 1965* | No | Yes | Yes | Yes | No | Yes | Yes | Yes |
| Plague: Shasta County, California  *United States Centres for Disease Control and Prevention, 1965* | No | Yes | Yes | Yes | No | Unclear | N/A | Yes |
| Suspected Case of Imported Bubonic Plague  *United States Centres for Disease Control and Prevention, 1966* | No | Yes | No | Yes | No | Unclear | N/A | Yes |
| Plague -- Arizona  *United States Centres for Disease Control and Prevention, 1967* | No | Yes | Yes | Yes | No | Unclear | N/A | Yes |
| Plague in San Diego  *Connor J. D., 1968* | Yes | Yes | Yes | Yes | No | N/A | N/A | Yes |
| Presumptive Bubonic Plague — Denver, Colorado  *United States Centres for Disease Control and Prevention, 1968* | No | Yes | Yes | Yes | No | Yes | Yes | Yes |
| Bubonic Plague Death -- Lemhi County, Idaho  *United States Centres for Disease Control and Prevention, 1968* | No | No | Yes | Yes | No | No | N/A | Yes |
| Plague Case -- Navajo Reservation -- Kayenta, Arizona  *United States Centres for Disease Control and Prevention, 1968* | No | Yes | Yes | Yes | No | No | Unclear | No |
| Plague -- New Mexico  *United States Centres for Disease Control and Prevention, 1969* | No | Yes | Yes | Yes | No | No | Unclear | No |
| Bubonic plague in the Southwestern United States  *Reed W. P., 1970* | No | Yes | Yes | Yes | No | Yes | Yes | Yes |
| Bubonic Plague – California  *United States Centres for Disease Control and Prevention, 1970* | No | Unclear | Unclear | Yes | No | No | N/A | No |
| Bubonic Plague – California  *United States Centres for Disease Control and Prevention, 1970* | No | Yes | Yes | Yes | No | No | N/A | Yes |
| Human Bubonic Plague -- Cochiti, New Mexico  *United States Centres for Disease Control and Prevention, 1970* | No | Yes | Yes | Yes | Yes | Yes | N/A | Yes |
| Bubonic Plague -- Santa Fe, New Mexico  *United States Centres for Disease Control and Prevention, 1970* | No | N/A | Yes | Yes | No | Unclear | N/A | Yes |
| Plague -- New Mexico  *United States Centres for Disease Control and Prevention, 1970* | No | Yes | Yes | Yes | No | Yes | N/A | Yes |
| Plague -- New Mexico  *United States Centres for Disease Control and Prevention, 1970* | No | Yes | Yes | Yes | No | No | N/A | Yes |
| Plague -- California  *United States Centres for Disease Control and Prevention, 1970* | No | Yes | Yes | Yes | No | No | N/A | Yes |
| Plague - Rio en Medio, New Mexico  *United States Centres for Disease Control and Prevention, 1970* | No | Yes | Yes | Yes | No | Unclear | N/A | Yes |
| Plague – New Mexico  *United States Centres for Disease Control and Prevention, 1970* | No | Yes | Yes | Yes | No | No | Unclear | No |
| Human Bubonic Plague – Oregon  *United States Centres for Disease Control and Prevention, 1971* | No | Yes | Yes | Yes | No | No | Unclear | No |
| Human Bubonic Plague – New Mexico  *United States Centres for Disease Control and Prevention, 1971* | No | Yes | Yes | Yes | No | Yes | N/A | Yes |
| Human Bubonic Plague - Coconino County Colorado  *United States Centres for Disease Control and Prevention, 1972* | No | Yes | Yes | Yes | No | No | N/A | Yes |
| Human Bubonic Plague – New Mexico  *United States Centres for Disease Control and Prevention, 1974* | No | Yes | Yes | No | N/A | N/A | N/A | Yes |
| Human Plague – New Mexico  *United States Centres for Disease Control and Prevention, 1974* | No | Yes | Yes | Yes | No | No | N/A | Yes |
| Human Plague – New Mexico, Utah  *United States Centres for Disease Control and Prevention, 1974* | No | Yes | Yes | Yes | No | No | N/A | Yes |
| Plague and the gallium scan  *Stahly T. L., 1975* | No | Yes | Yes | Yes | No | Yes | Yes | Yes |
| Fatal Human Plague – California  *United States Centres for Disease Control and Prevention, 1975* | No | Yes | Yes | Yes | No | Yes | N/A | Yes |
| Bubonic Plague – Arizona  *United States Centres for Disease Control and Prevention, 1975* | No | No | Yes | No | No | No | Unclear | Yes |
| Plague in Humans – New Mexico  *United States Centres for Disease Control and Prevention, 1975* | No | No | No | Yes | No | No | No | N/A |
| Human Plague Case -- Bernalillo County, New Mexico  *United States Centres for Disease Control and Prevention, 1975* | No | Yes | Yes | Yes | Yes | Yes | Yes | Yes |
| Bubonic Plague from Exposure to a Rabbit: A Documented Case, and a Review of Rabbit-Associated Plague Cases in The United States  *Von Reyn C. F., 1976* | No | Yes | Yes | Yes | No | Yes | N/A | Yes |
| Human Plague -- Arizona, California, New Mexico  *United States Centres for Disease Control and Prevention, 1976* | No | Yes | Yes | Yes | No | Unclear | No | Yes |
| Plague and pregnancy. A case report  *Mann J, 1977* | Yes | Yes | Yes | Yes | Yes | Yes | N/A | Yes |
| Plague -- Arizona, Colorado, New Mexico  *United States Centres for Disease Control and Prevention, 1977* | No | Yes | Yes | Yes | No | No | Yes | Yes |
| Plague – United States  *United States Centres for Disease Control and Prevention, 1977* | No | Yes | Yes | Yes | No | Unclear | N/A | Yes |
| Plague -- Arizona, California, New Mexico  *United States Centres for Disease Control and Prevention, 1978* | No | Yes | Yes | Yes | No | Yes | N/A | Yes |
| Plague in the United States: the "black death" is still alive  *Hoffman S. L., 1980* | No | Yes | Yes | Yes | No | Yes | N/A | Yes |
| Plague -- United States  *United States Centres for Disease Control and Prevention, 1980* | No | Yes | Yes | Yes | Yes | Yes | N/A | Yes |
| Human Plague – Texas, New Mexico  *United States Centres for Disease Control and Prevention, 1981* | No | Yes | Yes | Yes | No | Yes | Yes | Yes |
| Human plague associated with domestic cats--California, Colorado  *United States Centres for Disease Control and Prevention, 1981* | No | Yes | Yes | Yes | Yes | Yes | Yes | Yes |
| Peripatetic Plague  *Mann J., 1982* | No | Yes | Yes | Yes | Yes | Yes | N/A | Yes |
| Plague - South Carolina  *United States Centres for Disease Control and Prevention, 1983* | No | Yes | Yes | Yes | No | Yes | No | Yes |
| Plague Pneumonia – California  *United States Centres for Disease Control and Prevention, 1984* | No | Yes | No | Yes | No | No | No | Yes |
| Winter Plague -- Colorado, Washington, Texas, 1983-1984  *United States Centres for Disease Control and Prevention, 1984* | No | Yes | No | Yes | No | No | No | Yes |
| Human Bubonic Plague Transmitted by a Domestic Cat Scratch  *Weniger B. G., 1984* | No | Yes | Yes | Yes | No | Yes | N/A | Yes |
| Nineteen cases of plague in Arizona. A spectrum including ecthyma gangrenosum due to plague and plague in pregnancy  *Welty T. K., 1985* | No | Yes | Yes | Yes | No | Yes | Yes | Yes |
| Multiple lung cavities in a 12-year-old girl with bubonic plague, sepsis, and secondary pneumonia  *Florman A. L., 1986* | Yes | Yes | Yes | Yes | Yes | Yes | N/A | Yes |
| Plague in a pregnant patient  *Wong T. W., 1986* | No | Yes | Yes | Yes | Yes | Yes | N/A | Yes |
| Imaging in plague  *Moreno A. J., 1987* | No | No | Yes | Yes | No | Yes | Yes | Yes |
| Human Plague -- United States, 1988  *United States Centres for Disease Control and Prevention, 1988* | No | No | No | Unclear | No | No | No | No |
| Imported bubonic plague -- District of Columbia  *United States Centres for Disease Control and Prevention, 1990* | No | Yes | Yes | Yes | Yes | Yes | N/A | No |
| Bubonic plague in a child presenting with fever and altered mental status  *Migden D., 1990* | No | Yes | Yes | Yes | No | Yes | N/A | Yes |
| Plague in New Mexico  *Owens C., 1990* | No | Yes | Yes | Yes | No | No | N/A | Yes |
| An Outbreak of Plague in Northwestern Province, Zambia  *McClean K. L., 1995* | Yes | Yes | Yes | Yes | No | Yes | Yes | Yes |
| Fatal human plague--Arizona and Colorado, 1996  *United States Centres for Disease Control and Prevention, 1997* | No | Yes | Yes | Yes | N/A | N/A | N/A | Yes |
| Cases of cat-associated human plague in the Western US, 1977-1998  *Gage K. L., 2000* | No | Yes | Yes | Yes | No | No | Yes | Yes |
| Imported plague--New York City, 2002  *United States Centres for Disease Control and Prevention, 2003* | No | Yes | Yes | Yes | No | Yes | Yes | Yes |
| Painful lymphadenopathy and fulminant sepsis in a previously healthy 16-year-old girl  *Chmura K., 2003* | No | Yes | Yes | Yes | N/A | N/A | N/A | Yes |
| Human plague--four states, 2006  *United States Centres for Disease Control and Prevention, 2006* | No | Yes | No | Yes | No | No | No | Yes |
| Notes from the field: two cases of human plague--Oregon, 2010  *United States Centres for Disease Control and Prevention, 2011* | No | No | No | No | No | No | N/A | Yes |
| Misidentification of Yersinia pestis by Automated Systems, Resulting in Delayed Diagnoses of Human Plague Infections—Oregon and New Mexico, 2010–2011  *Tourdjman M., 2012* | No | Yes | Yes | Yes | No | Yes | Yes | Yes |
| Case report  *Lazet K., 2018* | No | Yes | Yes | Yes | Yes | Yes | N/A | Yes |
| Human case of bubonic plague resulting from the bite of a wild Gunnison’s prairie dog during translocation from a plague endemic area  *Melman S. D., 2018* | No | Yes | Yes | Yes | No | Yes | N/A | Yes |
| Two fatal cases of plague after consumption of raw marmot organs  *Kehrmann J., 2020* | Yes | Yes | Yes | Yes | Yes | Yes | Yes | Yes |
| Delays in Identification and Treatment of a Case of Septicemic Plague — Navajo County, Arizona, 2020  *Dale A. P., 2021* | Yes | Yes | Yes | Yes | Yes | Yes | NA | Yes |

**S2b – Risk of bias assessment for quasi-experimental studies**

| **Study title**  ***First author, Year*** | **Clear cause and effect** | **Participants similar in comparisons** | **Participants receiving similar treatment** | **Control group** | **Multiple measurements of outcome** | **Follow-up complete** | **Outcomes measured in same way** | **Outcomes measured in reliable way** | **Appropriate statistical analysis used** |
| --- | --- | --- | --- | --- | --- | --- | --- | --- | --- |
| Streptomycin in Bubonic Plague  *Haddad C. H., 1948* | No | Unclear | Unclear | No | No | Unclear | Unclear | Unclear | N/A |
| Co-trimoxazole in Bubonic Plague  *Nguyen-Van-Ai, 1973* | No | N/A | N/A | No | Unclear | Unclear | N/A | Unclear | N/A |
| Yersinia pestis Infection in Vietnam. II; Quantitative Blood Cultures and Detection of Endotoxin in the Cerebrospinal Fluid of Patients with Meningitis  *Butler T, 1976* | Yes | No | Yes | No | No | Unclear | Yes | Yes | N/A |

**S2c – Risk of bias assessment for cohort studies**

| **Study title**  ***First author, Year*** | **Similarities between groups** | **Similarity of measurement exposures** | **Exposure measured reliably** | **Confounding factors identified** | **Strategies to manage confounders** | **Groups free of outcome at moment of exposure** | **Measurement of outcomes reliable** | **Sufficient follow-up time** | **Follow-up complete** | **Strategies to address incomplete follow-up** | **Appropriate statistical analysis** |
| --- | --- | --- | --- | --- | --- | --- | --- | --- | --- | --- | --- |
| Clinical Features of Plague in the United States: the 1969-1970 Epidemic  *Palmer D. L., 1971* | N/A | N/A | Yes | No | N/A | Unclear | Unclear | No | Unclear | No | N/A |
| Yersinia pestis Infection in Vietnam. I. Clinical and Hematologic Aspects  *Butler T., 1974* | N/A | N/A | Yes | No | N/A | Unclear | Yes | Yes | Unclear | N/A | Yes |
| Epidemiological and clinical features of an outbreak of bubonic plague in New Mexico  *Von Reyn C. F., 1977* | N/A | N/A | Yes | No | No | Unclear | Unclear | Unclear | Unclear | No | Yes |
| Plague in the United States 1982  *Barnes A. M., 1983* | N/A | N/A | Yes | No | N/A | Unclear | Yes | Yes | Yes | N/A | Yes |
| Plague meningitis--a retrospective analysis of cases reported in the United States, 1970-1979  Becker T. M., 1987 | Unclear | Yes | Yes | No | N/A | N/A | Yes | N/A | N/A | N/A | Yes |
| Plague - A clinical review of 27 cases  *Crook L. D., 1992* | N/A | No | Yes | No | No | Unclear | Yes | No | Unclear | No | N/A |
| Current epidemiology of human plague in Madagascar  *Chanteau S., 2000* | Unclear | N/A | N/A | No | N/A | No | Yes | N/A | N/A | N/A | Yes |
| Gentamicin and Tetracyclines for the Treatment of Human Plague: Review of 75 cases in New Mexico, 1985-1999  *Boulanger L. L., 2004* | N/A | N/A | Yes | No | N/A | No | Yes | Unclear | Unclear | No | Yes |
| Plague Outbreak in Libya, 2009, Unrelated to Plague in Algeria  *Cabanel N., 2013* | N/A | Yes | Yes | Yes | No | Yes | Yes | Yes | Yes | N/A | Yes |
| Outbreak of Plague in a High Malaria Endemic Region — Nyimba District, Zambia, March–May 2015  *Sinyange N., 2016* | N/A | N/A | Yes | No | No | Unclear | Unclear | Unclear | Unclear | Unclear | Yes |
| Successful Treatment of Human Plague with Oral Ciprofloxacin  *Apangu T., 2017* | Yes | Yes | Yes | No | N/A | No | No | Yes | Yes | N/A | N/A |

**S2d – Risk of bias assessment for randomised controlled trials**

| **Study title**  ***First author, Year*** | **True randomisation** | **Concealed allocation** | **Treatment groups similar at baseline** | **Participants blind to allocation** | **Those delivering treatment blind to allocation** | **Outcome assessors blind to allocation** | **Groups treated identically** | **Follow-up complete** | **Participants analysed in groups allocated at randomisation** | **Outcomes measured in same way** | **Outcomes measured in reliable way** | **Appropriate statistical analysis** | **Protocol deviations accounted for** |
| --- | --- | --- | --- | --- | --- | --- | --- | --- | --- | --- | --- | --- | --- |
| Treatment of Plague with Gentamicin or Doxycycline in a Randomized Clinical Trial in Tanzania  *Mwengee W., 2006* | Yes | Unclear | Yes | No | No | No | Yes | No | Yes | Yes | Yes | Yes | Yes |
