## Supplemental Table 2 for "A systematic review of the clinical profile of patients with bubonic plague and the outcome measures used in research settings"

**S1 Table – Other reported signs and symptoms at baseline and post-baseline**

|  | N (%) patients for whom sign or symptom was reported | | | |
| --- | --- | --- | --- | --- |
| Sign or symptom | Baseline | Sample size | Post-baseline | Sample size |
| Abdominal distension | - | 0 | 4 | 107 |
| Acute renal failure | 3/5 (60%) | 5 | 1 | 2 |
| Altered mental status | 33/84 (39%) | 84 | 0 | 0 |
| Anaemia | - | 0 | 1 | 1 |
| Anorexia | 8/52 (15%) | 52 | 1 | 1 |
| Anterior hemiblock | 1/22 (5%) | 22 | 0 | 0 |
| Anuria | 1/1 (100%) | 1 | 0 | 0 |
| Anxiety | 2/28 (7%) | 28 | 1 | 1 |
| Arthralgia | 2/10 (20%) | 10 | 0 | 0 |
| Atelectasis | - | 0 | 1 | 1 |
| Bilateral pulmonary infiltrates | 1/4 (25%) | 4 | 1 | 4 |
| Bloodshot eyes | 1/3 (33%) | 3 | 0 | 0 |
| Bloody sputum | 1/3 (33%) | 3 | 5 | 7 |
| Blurred vision | 2/2 (100%) | 2 | 0 | 0 |
| Bradycardia | 1/22 (5%) | 22 | 0 | 0 |
| Breathing difficulty | 1/2 (50%) | 2 | 3 | 17 |
| Bubo suppuration | - | 0 | 2 | 65 |
| Bundle branch block | 1/22 (5%) | 22 | 0 | 0 |
| Cardiac arrest | - | 0 | 1 | 2 |
| Cellulitis | - | 0 | 3 | 18 |
| Cerebral edema | - | 0 | 1 | 1 |
| Chest heaviness | - | 0 | 1 | 1 |
| Chest pain | 1/17 (6%) | 17 | 3 | 27 |
| Cholelthiasis | - | 0 | 1 | 47 |
| Coma | 2/25 (8%) | 25 | 0 | 0 |
| Confusion | 4/46 (9%) | 46 | 0 | 0 |
| Congested | 1/3 (33%) | 3 | 0 | 0 |
| Congestive heart failure | - | 0 | 1 | 25 |
| Conjunctivitis | 1/19 (5%) | 19 | 1 | 25 |
| Conjunctival suffusion | 1/2 (50%) | 2 | 1 | 2 |
| Contractions | 1/1 (100%) | 1 | 0 | 0 |
| Costovertebral angle tenderness | 1/47 (2%) | 47 | 0 | 0 |
| Cutaneous ecchymoses | - | 0 | 1 | 4 |
| Cutaneous edema | 1/15 (7%) | 15 | 0 | 0 |
| Cyanosis | 1/2 (50%) | 2 | 1 | 1 |
| Decreased breath sounds | 1/1 (100%) | 1 | 0 | 0 |
| Decreased platelet count | - | 0 | 1 | 15 |
| Decreased white cell count | - | 0 | 1 | 25 |
| Decreased hematocrit | 1/1 (100%) | 1 | 0 | 0 |
| Dehydration | 4/4 (100%) | 4 | 0 | 0 |
| Delirium | 14/220 (6%) | 220 | 1 | 3 |
| Diaphoresis | 1/2 (50%) | 2 | 0 | 0 |
| Diarrhoea | 22/147 (15%) | 147 | 3 | 67 |
| Difficulty walking | 1/9 (11%) | 9 | 0 | 0 |
| Diffuse aching | 1/25 (4%) | 25 | 0 | 0 |
| Disorientation | 2/2 (100%) | 2 | 0 | 0 |
| Disseminated intravascular coagulation | 1/1 (100%) | 1 | 7 | 41 |
| Diverticulosis | - | 0 | 1 | 2 |
| Dizziness | 6/93 (6%) | 93 | 1 | 65 |
| Dry, coated tongue | 2/3 (67%) | 3 | 0 | 0 |
| Dyspnea | 2/10 (20%) | 10 | 2 | 6 |
| Ecthyma gangrenosum | 1/15 (6%) | 15 | 0 | 0 |
| Edema | - | 0 | 3 | 29 |
| Elevated blood pressure | 1/1 (100%) | 1 | 0 | 0 |
| Elevated liver function tests | 1/1 (100%) | 1 | 0 | 0 |
| Elevated transaminases | 1/1 (100%) | 2 | 0 | 0 |
| Elevated white cell count | 2/2 (100%) | 2 | 1 | 25 |
| Emaciation | - | 0 | 1 | 1 |
| Endophthalmitis | - | 0 | 1 | 1 |
| Endotoxemia | - | 0 | 3 | 42 |
| Enlarged liver | 1/2 (50%) | 2 | 0 | 0 |
| Erythema | 7/16 (44%) | 16 | 4 | 4 |
| Exophthalmos | - | 0 | 1 | 1 |
| Fetal distress | - | 0 | 1 | 1 |
| Fetal tachycardia | 1/1 (100%) | 1 | 1 | 1 |
| Fibrin thrombosis | - | 0 | 1 | 1 |
| Fluctuant nodes | - | 0 | 1 | 1 |
| Fluid filled lesion on thumb | 1/3 (33%) | 3 | 0 | 0 |
| Flushing | 1/1 (100%) | 1 | 1 | 1 |
| Fremitus | 1/1 (100%) | 1 | 0 | 0 |
| Gangrene | 1/1 (100%) | 1 | 0 | 0 |
| Gangrenous lymph node | 1/1 (100%) | 1 | 0 | 0 |
| Gastrointestinal bleeding | - | 0 | 1 | 17 |
| Granular casts | 1/1 (100%) | 1 | 0 | 0 |
| Hallucinations | 2/27 (7%) | 27 | 2 | 16 |
| Haemhorrhage - bilateral adrenal | - | 0 | 1 | 1 |
| Haemorrhage - ear | - | 0 | 1 | 1 |
| Haemorrhage - eye | - | 0 | 1 | 1 |
| Haemorrhage - lymph node | - | 0 | 1 | 25 |
| Hematuria | 1/1 (100%) | 1 | 0 | 0 |
| Hepatosplenomegaly | 1/1 (100%) | 1 | 1 | 1 |
| Herpes labialis | - | 0 | 1 | 65 |
| High proportion of immature neutrophils | 1/17 (6%) | 17 | 0 | 0 |
| Hyperventilation | - | 0 | 1 | 15 |
| Hypoactive blowel sounds | 1/47 (2%) | 47 | 0 | 0 |
| Hypotension | 15/87 (17%) | 87 | 11/85 (13%) | 85 |
| Hypoxemia with bilateral pulmonary edema | - | 0 | 1 | 1 |
| Icterus | - | 0 | 1 | 1 |
| Impaired consciousness | 2/2 (100%) | 2 | 1 | 3 |
| Incontinence | - | 0 | 1 | 3 |
| Increased partial thromboplastin times | - | 0 | 2 | 19 |
| Increased prothrombin time | - | 0 | 1 | 4 |
| Infectious syndrome | 1/2 (50%) | 2 | 0 | 0 |
| Inflammation of breast | 1/25 (4%) | 25 | 0 | 0 |
| Inflammation of shoulder | 1/25 (4%) | 25 | 0 | 0 |
| Injected conjunctivae | 1/3 (33%) | 3 | 0 | 0 |
| Inflamed eardrum | 1/1 (100%) | 1 | 0 | 0 |
| Inflamed throat | 1/1 (100%) | 1 | 0 | 0 |
| Irritability | 1/1 (100%) | 1 | 0 | 0 |
| Ischaemia | - | 0 | 1 | 2 |
| Jaundice | 1/3 (33%) | 3 | 0 | 0 |
| Lactic acid acidosis | - | 0 | 1 | 17 |
| Lesion haemorrhage on toe | 1/1 (100%) | 1 | 0 | 0 |
| Lethargy | 3/67 (4%) | 67 | 2 | 2 |
| Leukocytosis | 1/17 (6%) | 17 | 0 | 0 |
| Loss of appetite | 1/1 (100%) | 1 | 0 | 0 |
| Loss of consciousness | 1/1 (100%) | 1 | 0 | 0 |
| Lymph node necrosis | 1/4 (25%) | 4 | 1 | 25 |
| Lymphadenitis | - | 0 | 2 | 2 |
| Lymphadenopathy - generalised | - | 0 | 1 | 1 |
| Malaise | 22/68 (32%) | 68 | 1 | 1 |
| Membrane rupture | 1/1 (100%) | 1 | 0 | 0 |
| Meningeal irritation | - | 0 | 1 | 1 |
| Meningitis | 2/8 (25%) | 8 | 6 | 62 |
| Meningism | 2/28 (7%) | 28 | 0 | 0 |
| Metastatic anterior chamber endophthalmitis | 1/1 (100%) | 1 | 0 | 0 |
| Microhaematuria | 1/1 (100%) | 1 | 0 | 0 |
| Moribund appearance | - | 0 | 1 | 1 |
| Muscle pain | 2/26 (8%) | 26 | 0 | 0 |
| Myalgia | 54/204 (26%) | 204 | 0 | 0 |
| Myocarditis | - | 0 | 1 | 25 |
| Necrosis of the spleen | - | 0 | 1 | 1 |
| Non-purposeful movement of all extremities | 1/1 (100%) | 1 | 0 | 0 |
| Nuchal rigidity | 4/20 (20%) | 20 | 1 | 1 |
| Numbness and tingling sensation in arms | 1/1 (100%) | 1 | 0 | 0 |
| Obtunded | 2/77 (3%) | 77 | 1 | 15 |
| Osteomyelitis | - | 0 | 1 | 3 |
| Pain non-specific | 2/2 (100%) | 2 | 0 | 0 |
| Pain in arm | 3/17 (18%) | 17 | 0 | 0 |
| Pain in back | 2/18 (11%) | 18 | 0 | 0 |
| Pain in elbow | 1/1 (100%) | 1 | 0 | 0 |
| Pain in extremities | 1 | 1 | 0 | 0 |
| Pain in flank | 1 | 47 | 0 | 0 |
| Pain in groin | 1 | 2 | 1 | 1 |
| Pain at injection site | 0 | 0 | 1 | 1 |
| Pain in knee/leg | 1 | 1 | 0 | 0 |
| Pain in neck | 6 | 5 | 0 | 0 |
| Perianal abrasions | 1 | 1 | 0 | 0 |
| Pericarditis | 0 | 0 | 1 | 25 |
| Perivenous haemhorrage | 0 | 0 | 1 | 1 |
| Petechiae | 1 | 15 | 1 | 2 |
| Pharyngeal erythema | 3 | 8 | 0 | 0 |
| Photophobia | 1 | 25 | 0 | 0 |
| Pleocytosis | 0 | 0 | 1 | 1 |
| Pleural effusion | 0 | 0 | 2 | 5 |
| Pneumonia | 2 | 22 | 1 | 42 |
| Pneumonitis | 2 | 2 | 1 | 3 |
| Prostration | 3 | 21 | 1 | 1 |
| Proteinuria | 2 | 2 | 0 | 0 |
| Pulmonary edema | 1 | 1 | 1 | 1 |
| Pustular eruption | 0 | 0 | 1 | 1 |
| Pyuria | 2 | 2 | 0 | 0 |
| Rapid and shallow breathing | 2 | 4 | 1 | 1 |
| Rapid pulse | 0 | 0 | 1 | 1 |
| Rales | 1 | 1 | 0 | 0 |
| Rash | 2 | 4 | 1 | 1 |
| Rectal bleeding | 0 | 0 | 1 | 1 |
| Red blood cell casts | 1 | 1 | 0 | 0 |
| Red throat (with exudate on tonsils) | 2 | 2 | 0 | 0 |
| Reduced urine output | 0 | 0 | 1 | 1 |
| Refractory shock | 0 | 0 | 1 | 2 |
| Respiratory arrest/ failure | 1 | 1 | 3 | 19 |
| Respiratory distress | 2 | 4 | 5 | 46 |
| Restlessness | 1 | 3 | 0 | 0 |
| Rigors | 5 | 6 | 0 | 0 |
| Seizure | 4/34 (12%) | 34 | 2 | 66 |
| Septic shock | 5 | 59 | 2 | 4 |
| Skin lesions | 1 | 1 | 0 | 0 |
| Sinus tachycardia | 3 | 22 | 0 | 0 |
| Splenomegaly | 1 | 1 | 0 | 0 |
| ST-segment depressions | 1 | 22 | 0 | 0 |
| Staggering gait | 1 | 1 | 0 | 0 |
| Stiff back | 1 | 25 | 0 | 0 |
| Stiff neck | 2 | 26 | 1 | 3 |
| Stupor | 1 | 1 | 0 | 0 |
| Subpleural pulmonary haemorrhages | 0 | 0 | 1 | 1 |
| Sweating | 2 | 26 | 0 | 0 |
| Tachycardia | 4 | 28 | 1 | 25 |
| Tachypnoea | 1 | 1 | 2 | 2 |
| Thrombocytopenia | 2 | 2 | 4 | 25 |
| Thrombosis | 0 | 0 | 1 | 1 |
| Tonsillitis | 2 | 2 | 0 | 0 |
| Tracheal displacement | 1 | 4 | 0 | 0 |
| Tremor | 1 | 1 | 0 | 0 |
| Tubercule-like bodies in iris | 0 | 0 | 1 | 1 |
| Unspecified gastrointestinal complaints | 40 | 42 | 0 | 0 |
| Upper respiratory infection | 0 | 0 | 1 | 65 |
| Vertigo | 1 | 1 | 0 | 0 |
