## Supplemental Text 1 for "A systematic review of the clinical profile of patients with bubonic plague and the outcome measures used in research settings"

All searches were conducted in September 2021

**PubMED**

*Search terms:* ((("Bubonic plague"[Mesh]) OR (((plague[Title/Abstract]) OR "pasturella pestis"[Title/Abstract]) OR "yersinia pestis"[Title/Abstract]))) NOT (((("Animals"[Mesh] NOT ("Animals"[Mesh] AND "Humans"[Mesh])))))

*Article type*: case reports; clinical study; clinical trial; comparative study; controlled clinical trial; multicenter study; observational study; pragmatic clinical trial; randomised controlled trial

*Species*: humans

**JSTOR**

*Search terms*: plague OR "pasteurella pestis" OR "Yersinia pestis"

*Item type*: articles

*Search within results*: (patient* OR case* OR participant* OR subject* OR infant* OR child* OR adult*)

**Cochrane CENTRAL**

*Search terms*: ((("Plague"[Mesh]) OR (((plague[Title/Abstract]) OR "pasteurella pestis"[Title/Abstract]) OR "yersinia pestis"[Title/Abstract]))) NOT (((("Animals"[Mesh] NOT ("Animals"[Mesh] AND "Humans"[Mesh])))))

**International Clinical Trial Registry Platform (ICTRP)**

*Search term*: plague

**ISRCTN**

*Search term*: plague

**Clinicaltrials.gov**

*Search term*: plague
