## Supplementary figures and images for "A systematic review of the clinical profile of patients with bubonic plague and the outcome measures used in research settings"

### Supplemental Figure 1

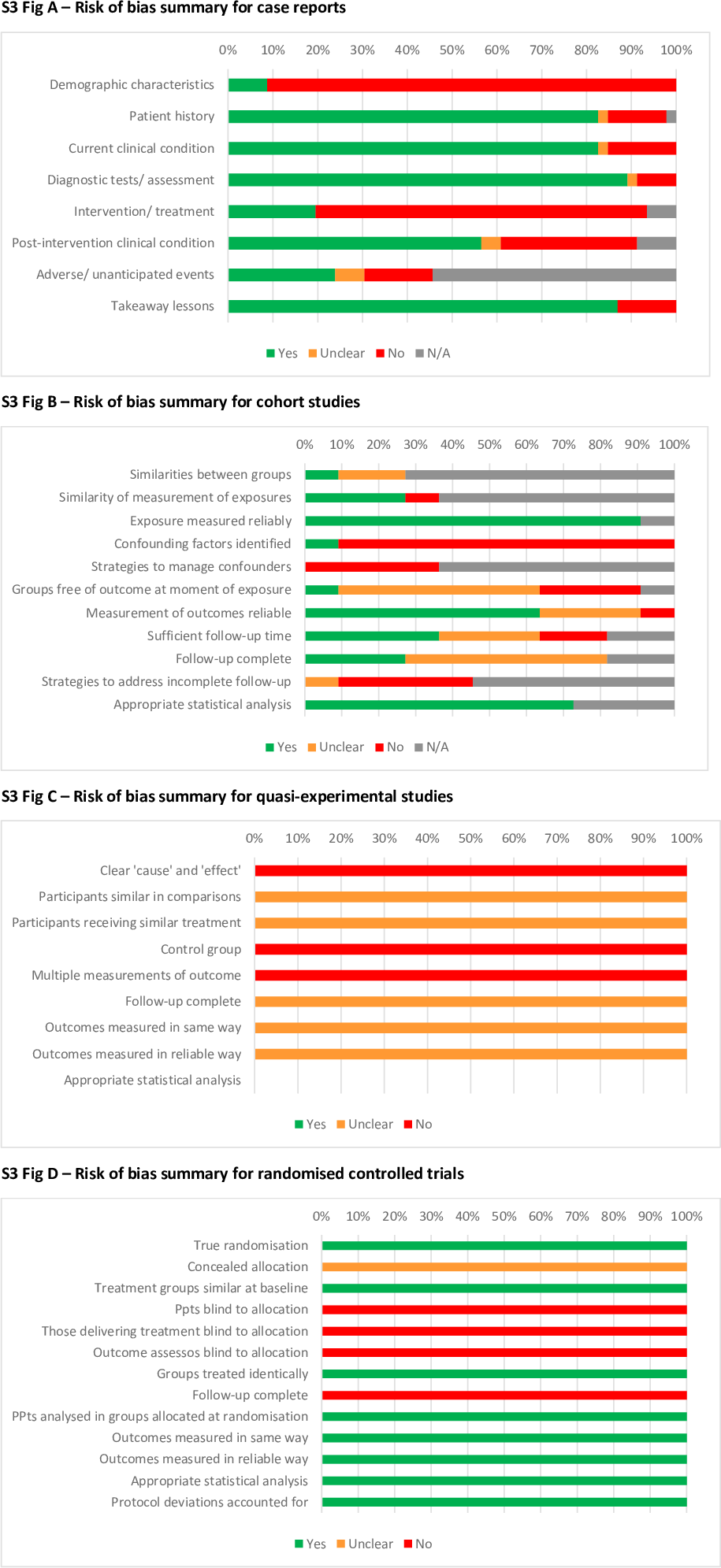
